# Persistent *Mycobacterium tuberculosis* bioaerosol release in a tuberculosis-endemic setting

**DOI:** 10.1101/2024.04.02.24305196

**Authors:** Ryan Dinkele, Sophia Gessner, Benjamin Patterson, Andrea McKerry, Zeenat Hoosen, Andiswa Vazi, Ronnett Seldon, Anastasia Koch, Digby F. Warner, Robin Wood

**Affiliations:** UCT Molecular Mycobacteriology Research Unit, Department of Pathology, Faculty of Health Sciences, University of Cape Town, Cape Town, 7925, South Africa; Institute of Infectious Disease and Molecular Medicine, Faculty of Health Sciences, University of Cape Town, Cape Town, 7925, South Africa; Amsterdam Institute for Global Health and Development, University of Amsterdam, Amsterdam, 1105, The Netherlands; Aerobiology and TB Research Unit, Desmond Tutu Health Foundation, Cape Town, 7925, South Africa; Wellcome Centre for Infectious Diseases Research in Africa, Faculty of Health Sciences, University of Cape Town, Cape Town, 7925, South Africa

## Abstract

Pioneering studies linking symptomatic disease and cough-mediated release of *Mycobacterium tuberculosis* (*Mtb*) established the infectious origin of tuberculosis (TB), simultaneously informing the pervasive notion that pathology is a prerequisite for *Mtb* transmission. Our prior work has challenged this assumption: by sampling TB clinic attendees, we detected equivalent release of *Mtb*-containing bioaerosols by confirmed TB patients and individuals not receiving a TB diagnosis, and we demonstrated a time-dependent reduction in *Mtb* bioaerosol positivity during six-months’ follow-up, irrespective of anti-TB chemotherapy. Now, by extending bioaerosol sampling to a randomly selected community cohort, we show that *Mtb* release is common in a TB-endemic setting: of 89 participants, 79.8% (71/89) produced *Mtb* bioaerosols independently of QuantiFERON-TB Gold status, a standard test for *Mtb* infection; moreover, during two-months’ longitudinal sampling, only 2% (1/50) were serially *Mtb* bioaerosol negative. These results necessitate a reframing of the prevailing paradigm of *Mtb* transmission and infection, and may explain the current inability to elucidate *Mtb* transmission networks in TB-endemic regions.

**Summary:** Elucidating chains of *Mycobacterium tuberculosis* transmission is limited by a dependence on linking sputum-positive tuberculosis cases. Here, we report persistent *M. tuberculosis* bioaerosol release in the majority of a randomly selected community cohort. The contribution to tuberculosis transmission is unknown.

## Introduction

Infection with *Mycobacterium tuberculosis* (*Mtb*) encompasses a spectrum of outcomes (Drain et al., 2018, Barry et al., 2009, Lin and Flynn, 2018, Coussens et al., 2024). At its mildest, the immune system clears or contains infecting *Mtb* bacilli with minimal discernible impact on the host (Richards et al., 2023). At its most severe, pulmonary *Mtb* infection results in advanced disease that is associated with a 40–70% fatality rate in the absence of effective treatment (Hermans et al., 2015). Despite the rarity of progression to active disease at an individual level and the availability of effective chemotherapy (Richards et al., 2023, Horton et al., 2023), tuberculosis (TB) remains a leading global cause of mortality (WHO, 2023), with notification rates surpassing those reported in the early 1900s in certain settings (Hermans et al., 2015).

In endemic regions, most incident TB is thought to arise from the recent (<2 years) transmission of *Mtb* (Shah et al., 2017, Dowdy and Behr, 2022). However, with only 1–30% of new *Mtb* infections traceable to known TB cases (Crampin et al., 2006, Glynn et al., 2015, Middelkoop et al., 2015, Verver et al., 2004), it is likely that numerous undetected sources of *Mtb* transmission exist within these communities. Consistent with this possibility, there is growing interest in the notion that early TB states – encompassing *Mtb* infection, subclinical, and clinical TB (Coussens et al., 2024) – might provide a previously overlooked reservoir of transmission (Kendall et al., 2021, Nguyen et al., 2023). Establishing the propensity for *Mtb* release – often equated with “infectiousness” – during early TB is therefore essential, but very difficult to accomplish (Dinkele et al., 2024).

Sputum diagnosis is generally considered the gold standard for assessing infectiousness among TB patients (Pai et al., 2023). Through this lens, data from extensive community TB screening efforts utilizing sputum-GeneXpert – which have identified large numbers of asymptomatic cases (Moyo et al., 2022) – support the notion of unrecognized transmitters in high-burden communities. And, while these results accommodate the potential for *Mtb* transmission to occur prior to the development of recognizable TB symptoms (Nguyen et al., 2023), they nevertheless rely on sputum production (itself a symptom of disease) for the detection of *Mtb*. This dependency on sputum production imposes a significant constraint on attempts to detect individuals with early TB, and is exacerbated by the difficulties encountered in obtaining useable samples from many individuals (Pai et al., 2023).

Bioaerosol sampling offers a non-invasive method to collect peripheral lung fluid and/or particulate matter independent of the specific disease state (Johnson and Morawska, 2009). Moreover, the detection of *Mtb* in bioaerosols signifies the release of bacilli by an individual, tentatively offering a concurrent metric of transmission risk (Williams et al., 2023, Jones-López et al., 2013). Despite these advantages, the utilization of bioaerosol sampling has predominantly been restricted to individuals with bacteriologically confirmed TB. This owes mainly to the challenges inherent in handling extremely paucibacillary samples as well as the assumed importance of pathology for *Mtb* release – in turn reinforcing the primacy of sputum bacterial burden as key predictor of infectiousness. Insights gained from the COVID-19 pandemic, however, established the plausibility of asymptomatic transmission (Shaikh et al., 2023), with analogous recent results suggesting the cough-independent aerosolization of *Mtb* (Dinkele et al., 2022, Williams et al., 2020). It seems possible, therefore, that individuals might progress from *Mtb* infected (often inferred via interferon gamma release assay; IGRA (Cohen et al., 2019)) to infectious before producing sputum.

Since 2013, we have studied the spontaneous generation of *Mtb*-containing bioaerosols using the Respiratory Aerosol Sampling Chamber (RASC), a highly sensitive, adaptable personal clean room designed to capture all particulate matter (including *Mtb* bacilli) released by individuals (Dinkele et al., 2021). By combining liquid capture of bioaerosols in the RASC with DMN-trehalose-enabled microscopic detection and visualization of *Mtb* bacilli, we recently reported that ∼90% of clinic attendees with presumptive TB produced bioaerosols containing viable *Mtb* bacilli (Patterson et al., 2024). Surprisingly, the high proportion of *Mtb* bioaerosol-positivity occurred irrespective of final TB diagnosis and included both notified TB patients – who commenced anti-TB chemotherapy – and those not diagnosed with TB (who did not initiate treatment). Moreover, during six-months’ follow-up, the rates of decline in symptom severity and *Mtb* bioaerosol release were equivalent in both groups. And, at the six-month study endpoint, approximately 20% of all participants – including treated TB patients and those who did not receive a TB diagnosis – remained *Mtb* bioaerosol-positive despite clinical resolution, consistent with prior observations from radiological assays (Malherbe et al., 2016).

Given the high-prevalence of *Mtb* bioaerosol release among clinic attendees not diagnosed with TB, as well as the frequency of *Mtb* bioaerosol release in confirmed TB patients on completion of standard anti-TB treatment, we hypothesized that the production of *Mtb* bioaerosols by individuals living in TB-endemic communities might be more common than previously thought. To investigate this possibility, we utilized our bioaerosol sampling platform (Dinkele et al., 2022) to investigate *Mtb* release in 89 randomly selected participants recruited into two consecutive cohorts: the first was a cross-sectional community survey comprising 39 participants, and the second was a longitudinal observational study in which 50 individuals consented to be sampled at three separate timepoints over two months. As detailed below, our results suggest that early-stage *Mtb* infection is pervasive and may be an important contributor to the transmission of *Mtb* in high TB-burden communities.

## Results

### The high prevalence of Mtb bioaerosol release occurs independently of respiratory maneuver

To investigate the prevalence of *Mtb* bioaerosol release in a TB-endemic setting, we initiated a cross-sectional community survey in Masiphumelele, Cape Town, with recruitment randomized geospatially by parcel of land, or “erf” (Fig. 1). From 10 erfs, we sequentially enrolled 39 individuals (1–12/erf [median=3]) who were assessed in *Mtb* bioaerosol release, sputum-GeneXpert MTB/RIF (GXP), and QuantiFERON-TB Gold (QFT) assays. The cohort had high rates of Human Immunodeficiency Virus (HIV) infection (13%) and previous TB (18%). Two participants who produced *Mtb* bioaerosols at the screening visit (5%) were GXP-positive at follow-up (one of these participants had completed treatment for TB in 2020, two years prior to the study), providing immediate evidence of undiagnosed TB in the community (Table 1).

**Table 1:** Summary of participant demographic and clinical information from the two cohorts.

| Variable | Category | Cohort 1 | Cohort 2 | P value |
| --- | --- | --- | --- | --- |
|  |  | n (%) |  |  |
| Total |  | 39 (100) | 50 (100) | - |
| Gender | Male | 12 (30.8) | 13 (26) | 0.64 |
| Previous TB | Yes | 7 (17.9) | 4 (8) | 0.20 |
| HIV status | Negative | 29 (74.4) | 38 (76) | **0.02** |
|  | Positive | 5 (12.8) | 12 (24) |  |
|  | Unknown | 5 (12.8) | 0 (0) |  |
| GeneXpert Ultra | Negative | 34 (87.2) | 48 (96) | >0.99 |
|  | Positive | 2 (5.1) | 2 (4) |  |
|  | NA* | 3 (7.7) | 0 (0) |  |
| QuantiFERON-TB Gold | Negative | 17 (43.6) | 16 (32) | 0.15 |
|  | Positive | 16 (41) | 33 (66) |  |
|  | Indeterminate | 2 (5.1) | 1 (2) |  |
|  | NA* | 4 (10.3) | 0 (0) |  |
| Age [mean (sd)] |  | 32.9 (12.6) | 34.2 (12.5) | 0.636 |

NA = Not available (Incomplete or failed assay), \*Excluded from the analysis

**Figure 1:**
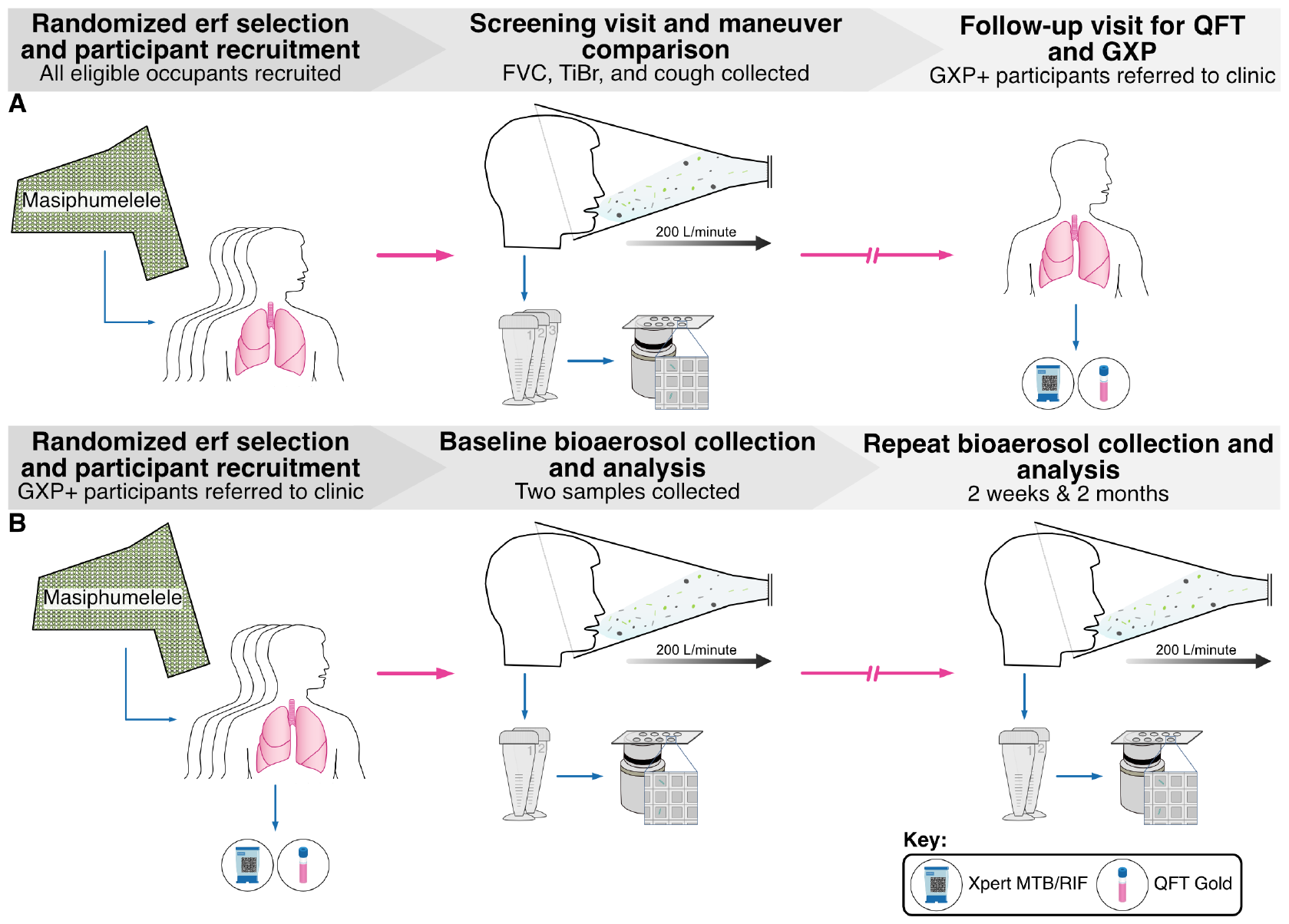
Participant recruitment and bioaerosol sampling algorithms for the two cohorts. (**A**) 39 participants were recruited from randomly selected erfs in Masiphumelele. At a first screening visit, participants produced bioaerosol samples from three respiratory maneuvers; forced vital capacity (FVC), tidal breathing (TiBr), and induced cough. Samples were processed and visualized independently by microscopists blinded to all sample information. Owing to the high prevalence of *Mtb* bioaerosol positivity, all participants were brought back for a follow-up visit during which blood and sputum were collected for QFT and GXP analysis, respectively. (**B**) 50 participants were recruited into a longitudinal observational study of *Mtb* release. Blood and sputum samples were collected at baseline for QFT and GXP analyses, respectively. Two equivalent bioaerosol samples were collected during ten minutes of tidal breathing with deep breaths taken at 30-second intervals. These samples were processed and imaged independently on nanowell-arrayed microscope slides by microscopists blinded to all sample information. This process was repeated at two weeks and two months post initial recruitment.

Advanced lung pathology and chronic cough are often viewed as essential for *Mtb* release (Turner and Bothamley, 2015). This assumption is, however, incompatible with the notion that *Mtb* may transmit during early-stage infection. Here, we examined the potential for *Mtb* release from a randomly selected community cohort, considering both unrelated cough (Esmail et al., 2018) and tidal breathing (Dinkele et al., 2022) as potential mechanisms. This was done by comparing *Mtb* aerosolization from three independently sampled respiratory maneuvers; namely, forced vital capacity (FVC – a deep breathing respiratory maneuver), tidal breathing (TiBr), and cough, according to our previously described methodology (Dinkele et al., 2022). When comparing the respiratory maneuvers, the percentage of *Mtb*-positive samples ranged from 51.1–67.6%, with no significant differences observed in the likelihood of producing *Mtb* between the three respiratory maneuvers (Fig. 2A & B). Moreover, no differences were observed in the average number of bacteria identified per sample (Fig. 2C & D). Across FVC, TiBr, and cough, the average counts were 2.9 (95% CI: 1.9; 4.0), 2.2 (95% CI: 1.4; 3.2), and 2.0 (95% CI: 1.1; 3.1) *Mtb* bacilli per sample, respectively. Overall, we found that 79.5% (31/39) of the participants produced at least one positive bioaerosol sample when looking across all three maneuvers. Together, these data suggest that the prevalence of *Mtb* release is high in a TB-endemic setting.

**Figure 2:**
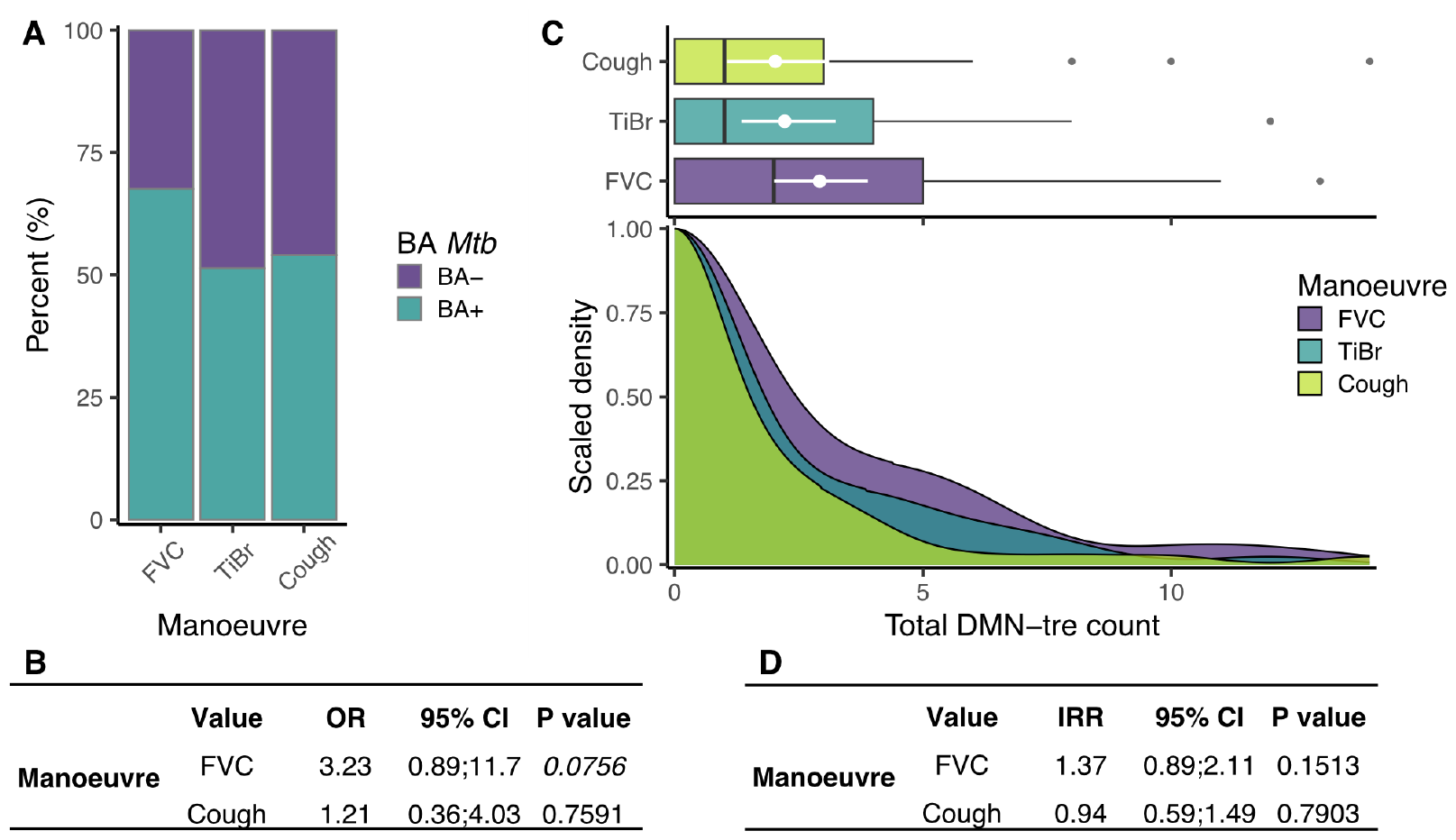
The production of aerosolized *Mtb* during all three respiratory maneuvers in the first cohort. (**A**) The percentage of samples in which putative *Mtb* were detected (turquoise) or absent (purple) from forced vital capacity [FVC (67.6%)], tidal breathing [TiBr (51.4%)], and cough (51.1%). (**B**) Results of a logistic regression comparing the odds of a positive bioaerosol result compared to TiBr. **(C)** Box and density plots comparing the total number of *Mtb* detected between the three respiratory maneuvers. (**D**) Results of a negative binomial regression comparing the number of *Mtb* detected between the three respiratory maneuvers. OR = odds ratio, IRR = incident rate ratio, CI = confidence interval, BA = bioaerosol.

### Altering the bioaerosol sampling algorithm did not reduce Mtb detection efficiency

The observation that ∼80% of a randomly selected community cohort produced *Mtb* bioaerosols was reminiscent of our previous work, which demonstrated the high (∼90%) prevalence of *Mtb* release among clinic attendees with presumed TB, regardless of final diagnosis or respiratory maneuver (Patterson et al., 2024). However, the cross-sectional community survey only provided a snapshot in time of *Mtb* bioaerosol release. To investigate the variability of *Mtb* release through time, we repeated our random community sampling strategy in recruiting a further 50 individuals from 20 erfs (1–7/erf [median=2]) into a longitudinal observational study of *Mtb* release (Fig. 1).

Bioaerosols were collected at baseline, and follow-up visits were scheduled at two weeks and two months post baseline, in accordance with previously described time intervals (Patterson et al., 2024). The characteristics of this cohort were not dissimilar to the first 39 participants (Table 1), except for HIV status – where the difference was driven largely by the proportion of individuals reporting their status as “unknown”.

Given the observation that FVC and TiBr were sufficient to aerosolize *Mtb*, we reasoned our bioaerosol sampling algorithm could be simplified by removing induced cough. Therefore, rather than collecting three five-minute samples (∼15 minutes), each from a different respiratory maneuver, we implemented a single sampling algorithm which was repeated twice: ten minutes of tidal breathing with deep breaths every 30 seconds (∼20 minutes sampling in total). This enabled a head-to-head comparison of duplicate bioaerosol samples at a single visit and allowed for an assessment of variation through time. We found comparable results between first and second cohorts in both the proportion of participants producing at least one positive sample and the total number of aerosolized *Mtb* (Fig. 3A–C). Average *Mtb* counts at baseline varied from 0– 16, with 20% (10/50) of the participants producing two negative bioaerosol samples (Fig. 4A). Discrepant samples (*i*.*e*., where one sample was negative and the other positive), were relatively rare [22% (11/50)] with a low maximum sample difference (3 bacilli). There was a strong correlation between the first and second bioaerosol sample (Fig. 4B) and a high degree of agreement, with 60% of the samples differing by ± 1 (Fig. 4C & D). Considering the relatively small sample size of each cohort and the comparability of the bioaerosol results, we pooled the data to investigate which covariates were associated with *Mtb* release. No covariates were associated with increased odds of a positive sample (Table 2). In contrast, biological sex (IRR = 1.88, p < 0.05), previous TB (IRR = 2.33, p < 0.05), and age ≥45 (IRR = 0.48, p < 0.05) were associated with the number of *Mtb* detected (Table 3). Surprisingly, neither QFT status – the conventional marker of *Mtb* infection – nor HIV status were associated with the associated with *Mtb* release. Overall, these data suggest that individuals who aerosolize *Mtb* do so consistently within short time frames and independently of previously accepted markers of infection.

**Table 2:**
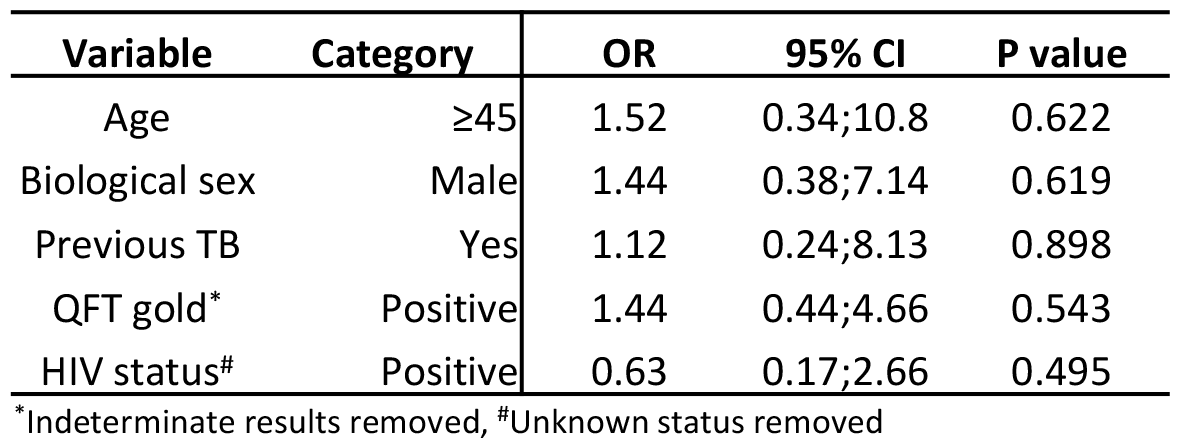
Results of a logistic regression assessing the odds of a positive bioaerosol sample. OR = odds ratio, CI = confidence interval.

**Table 3:**
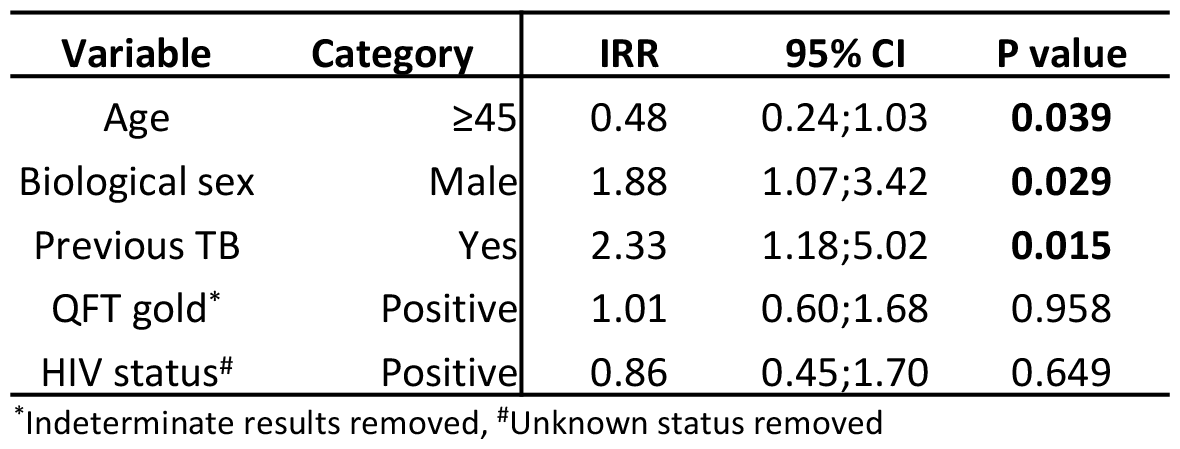
Results of a negative binomial regression assessing the number of *Mtb* per participant. IRR = incident rate ratio, CI = confidence interval.

**Figure 3:**
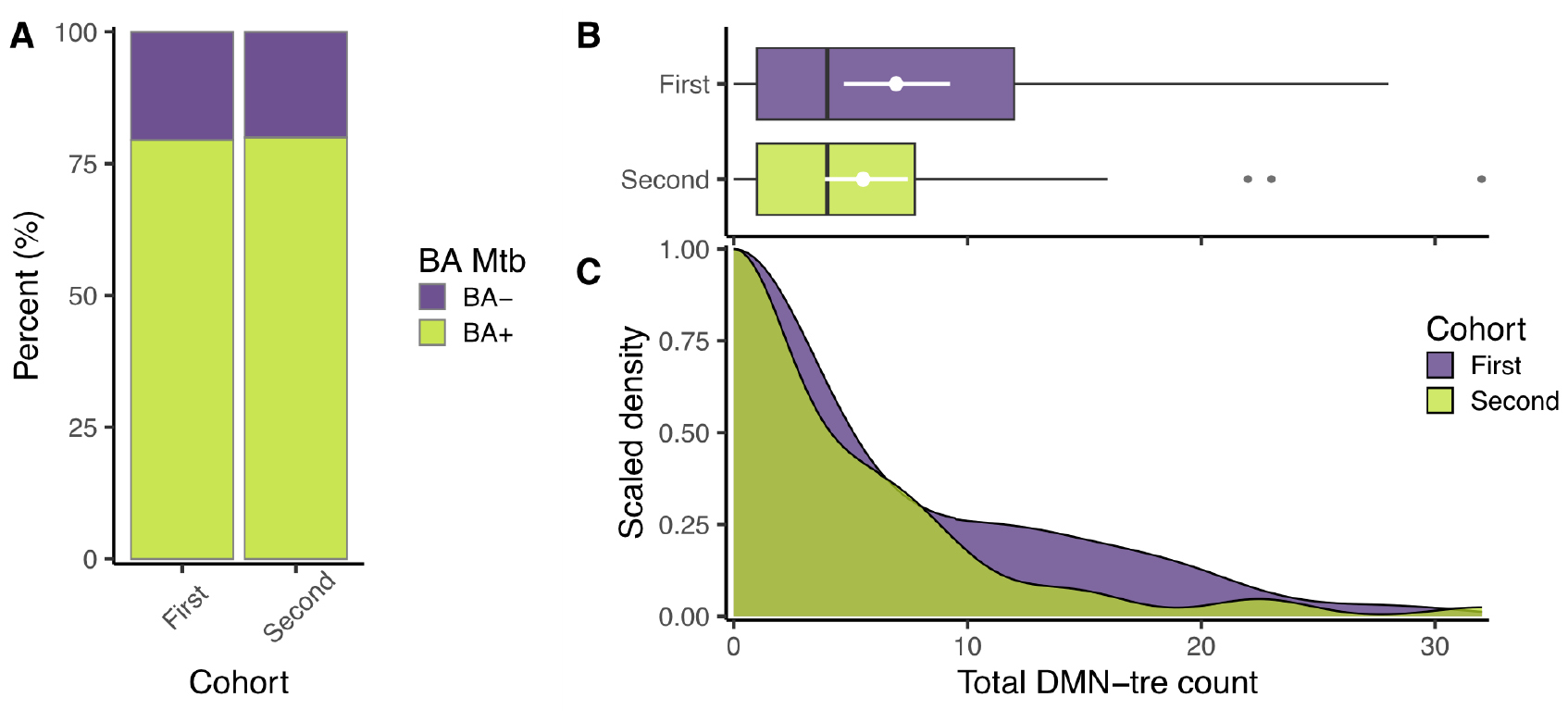
The production of aerosolized *Mtb* between the two cohorts. (**A**) The percentage of samples in which putative *Mtb* were detected (green) or absent (purple). The odds of a positive bioaerosol sample were equivalent between the two groups (OR = 1.03, 95% CI = 0.36;2.92, p = 0.952) (**B**) Box and whisker and (**C**) equivalent density plots comparing the total number of *Mtb* detected between the two cohorts. The rates at which *Mtb* were produced during the two samplings were equivalent (IRR = 0.797, 95% CI = 0.47;1.33, p = 0.386). OR = odds ratio, IRR = incident rate ratio, CI = confidence interval, BA = bioaerosol.

**Figure 4:**
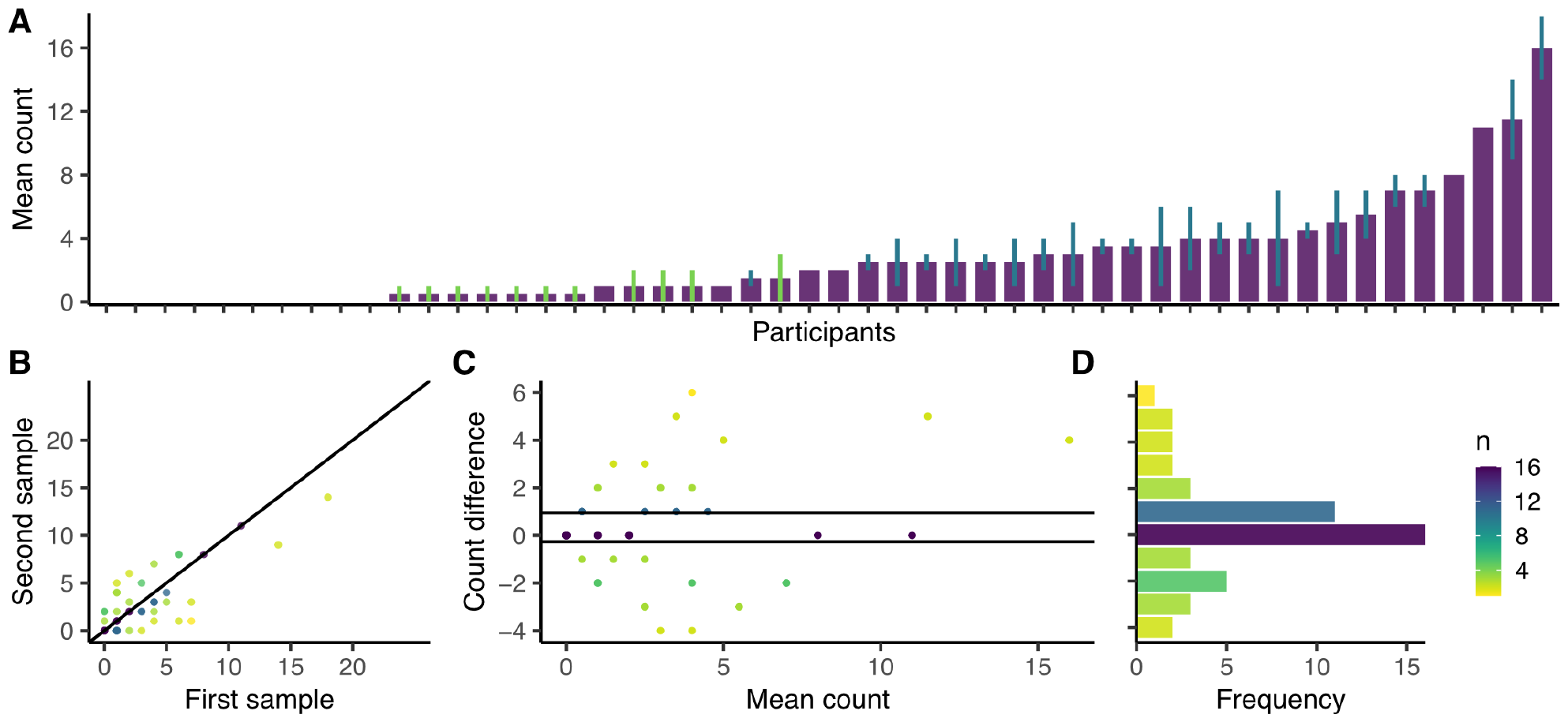
The production of aerosolized *Mtb* during two equivalent respiratory maneuvers in the second cohort. (**A**) Plot of the mean *Mtb* (DMN-tre positive) count from two samples, with error bars representing the range. No lines indicate equal counts, green lines indicate one count = 0, blue lines indicate two counts > 0. (**B**) Plot of the *Mtb* (DMN-tre positive) counts of the first and second samples at baseline (r = 0.810, p < 0.0001) with a fitted line representing a 1:1 correlation. (**C**) A Bland-Altman plot indicating the level of agreement between the first and second samples, with a (**D**) histogram showing the frequency of each count difference. Most samples differed by either 0 or ±1 (60%) and 94% of the samples differed by four or less.

### Persistent Mtb release among a randomly selected community cohort is common

Based on the observation from baseline sampling that 94% of matched samples differed by less than five, we reasoned that differences in the average count between two consecutive samples through time of five or greater could be considered significant. According to this definition, most [74% (32/43)] participants were constant low-level producers of *Mtb* over two months (Fig. 5A). The rate of bioaerosol positivity was relatively consistent through time, ranging from 76.7–90.7% (Fig. 5B & C). Most surprisingly, only one individual was negative across all three visits (representing six negative bioaerosol samples). Of 43 individuals with complete data for all three timepoints (baseline, two weeks, two months), 19% (8/43) and 9% (4/43) were negative at one and two visits, respectively. Although individual participants varied in their bioaerosol count from visit to visit, there was no overall trend in the number of *Mtb* detected per visit (Fig. 5D & E). This lack of secular trend in *Mtb* bioaerosol release among a randomly selected community cohort supports our previous hypotheses that *Mtb* bioaerosol clearance is immune driven (Patterson et al., 2024).

**Figure 5:**
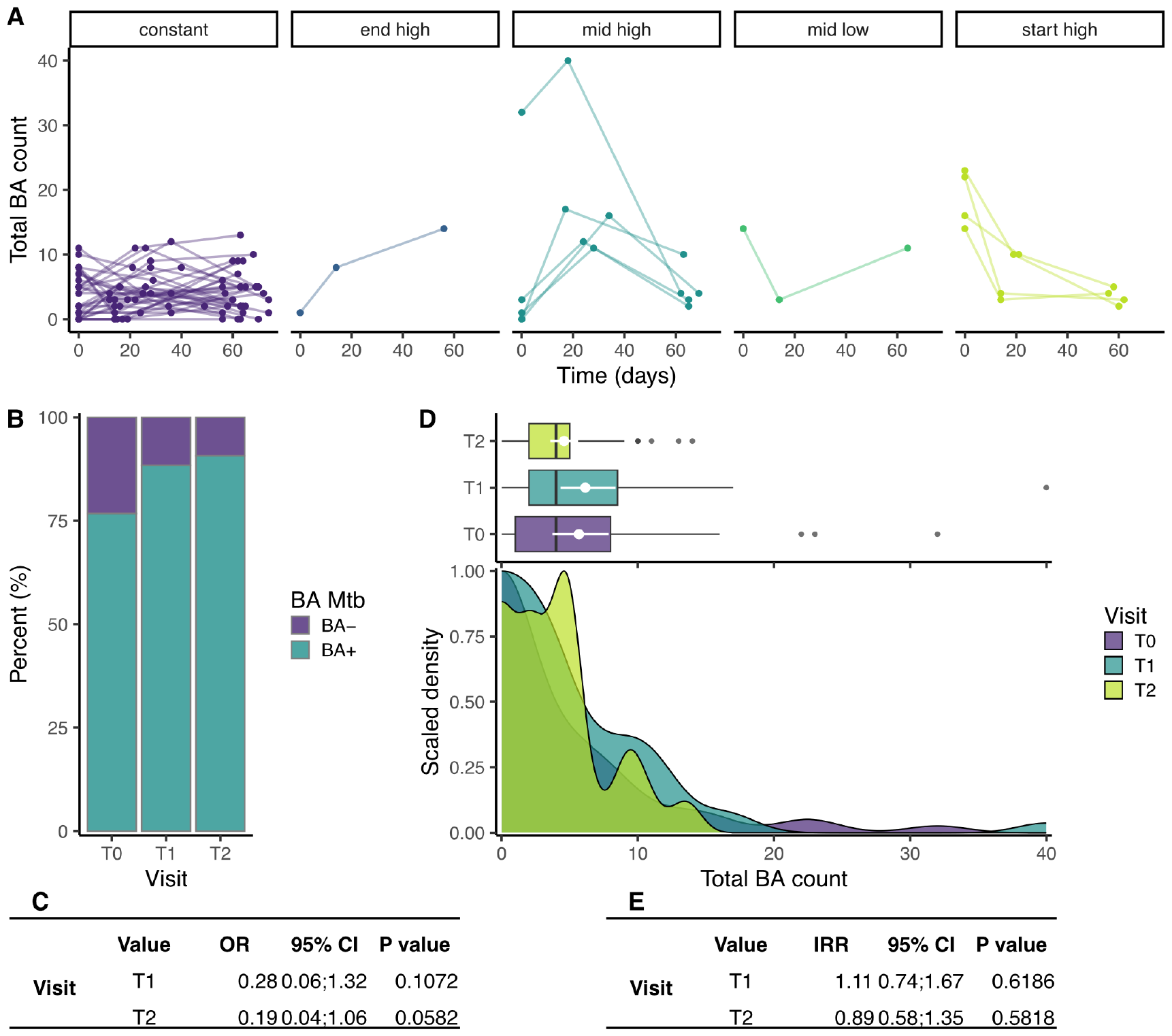
The production of aerosolized *Mtb* through time in the second cohort. (**A**) Total Mtb (DMN-tre positive) counts (sum of the two samples) through time, stratified by time trend. (**B**) The percentage of samples in which putative *Mtb* were detected (turquoise) or absent (purple) at each of the visits. (**C**) Results of a logistic regression comparing the odds of a negative BA result compared to T0. (**D**) Box and density plots comparing the total number of *Mtb* bacilli (DMN-tre positive) detected at each visit. (**E**) Results of a negative binomial regression comparing the number of *Mtb* bacilli (DMN-tre positive) detected at each visit. OR = odds ratio, IRR = incident rate ratio, CI = confidence interval, BA = bioaerosol.

## Discussion

The use of new tools in TB research has enabled unexpected insights that question established models of pathogenicity. For *Mtb* transmission, the increasing awareness that TB disease is not required for *Mtb* bioaerosol release interrogates previous assumptions linking pathology to infectiousness (Dinkele et al., 2024), in turn raising important questions about the implications for new diagnostics, drugs, and vaccines (Fortune, 2024).

In work immediately preceding this report, we detected high-prevalence *Mtb* bioaerosol release among TB clinic attendees diagnosed as not having TB, as well as the persistent release of *Mtb* bioaerosols post-treatment among bacteriologically confirmed TB cases (Patterson et al., 2024) – a finding consistent with separate work which used positron emission tomography–computed tomography (PET-CT) imaging and *Mtb* mRNA detection (Malherbe et al 2016) to conclude that apparently curative treatment for TB might not eradicate all *Mtb* bacteria. One implication of our previous results was that release of *Mtb* bioaerosols within TB-endemic communities might be considerably more prevalent than previously understood. To address that possibility, we initiated the work reported here, in which we broadened our sampled population to screen a random selection of individuals from the larger of the two communities served in the earlier study. In doing so, we modified the sampling protocol slightly to exclude unnecessary forced coughing and, importantly, demonstrated high reproducibility in *Mtb* capture numbers when repeated samples were obtained from the same individual at the same visit.

The results were again very unexpected: we identified aerosolized *Mtb* in 80% of participants recruited at random from a high TB-burden community. Moreover, *Mtb* release occurred independently of QFT results, the test conventionally used to detect current or previous exposure to *Mtb* (Cohen et al., 2019). Together, these observations suggest that current methods for detecting *Mtb* infection and estimating patient infectiousness are insufficient, and that *Mtb* release is far more common in TB-endemic settings than previous appreciated. They also imply a large reservoir of potential transmitters that remains invisible to national TB programs, although definitive demonstration of *Mtb* transmission from this population of relatively well individuals is necessary. Further studies are planned to establish the attributable fraction of TB transmission from this population; the immediate implications, though, are that a focus on treatment as the sole intervention to prevent transmission will require reassessing if the attributable fraction is significant in high TB-burdened settings. In addition, the potential utility of immune strategies including vaccination might be negatively impacted if these organisms are able to avoid immune surveillance or are in a relatively immune-privileged site.

Given the assumed importance of lung pathology and chronic cough in TB transmission (Turner and Bothamley, 2015), there are currently few mechanistic explanations for how bacilli are aerosolized during early-stage *Mtb* infection. Notably, from longitudinal sampling of 50 participants over 2 months, it was apparent that most individuals were transiently or persistently releasing *Mtb*, with only one individual returning negative *Mtb*-bioaerosol results across all three visits (representing six negative bioaerosol samples). Despite variation in *Mtb* release per participant, no overall trend was observed in *Mtb* release through time for the cohort. This observation supports our previous inference that, among a predominantly symptomatic cohort of clinic attendees with presumptive TB, the reduction in *Mtb* release through time might be largely immune driven (Patterson et al., 2024).

These results might offer new insights into the *Mtb* host-pathogen interaction. It is tempting to suggest, for example, that the persistent bioaerosol release of *Mtb* within this community could indicate a homeostatic interaction between the bacillus and its human host. Although half of the participants were QFT positive, there was no correlation with aerosol *Mtb* exhalation. This could be explained if the aerosolized *Mtb* phenotype remains immunologically unrecognized or is present only in a relatively immune-privileged site. Notably, HIV status was not associated with aerosolized *Mtb*, further implying a lack of immune interaction between host and pathogen. In this context, recent work suggesting intercellular replication of *Mtb* within biofilm-like cords might be apposite (Mishra et al., 2023). Determining the anatomical location of bacilli released in bioaerosols in asymptomatic individuals is a key research priority.

Our results must be interpreted in the light of the study’s limitations, including sample size and composition, as well as the potential for the false-positive identification of *Mtb*. Participants were recruited from randomly selected erfs during the workday between Monday–Friday. Our sample was therefore biased to individuals both willing and able to participate in the study. However, the average age, QFT status, and HIV prevalence were as expected for the community (Middelkoop et al., 2014), suggesting that our observations are internally reliable within Masiphumelele. It seems unlikely, therefore, that the manner of participant recruitment would significantly impact the results of this study.

Our assay for the detection of *Mtb* release is based on the microscopic detection of bacilli matching specific morphological characteristics, which also take up with DMN-tre (Dinkele et al., 2021). This technique is potentially subject to the false-positive identification of *Mtb*, given that other actinomycetales produce the enzyme required for the probe incorporation (Kamariza et al., 2018). However, we have previously used the RASC for *Mtb* detection by colony formation and droplet-digital PCR (ddPCR) (Wood et al., 2016, Patterson et al., 2018, Patterson et al., 2024). More recently, we successfully obtained whole-genome sequence data from three bioaerosol samples after fifty-day culture, importantly demonstrating the presence of *Mtb*, with no non-tubercular mycobacteria or other actinomycetales found (Patterson et al., 2024).

With these limitations in mind, we interpret our results as indicating that *Mtb* bioaerosol release is common in this community and is not detectable by standard measures of *Mtb* infection. If correct, these observations motivate for an urgent reframing of the prevailing paradigm of *Mtb* transmission and infection. The notion that infection with *Mtb* constitutes the primary determinant of TB risk in high-TB burden settings has dominated thinking about new TB interventions. Our work using advanced breath aerosol collection technology challenges this assumption: in successive studies, we have detected release of *Mtb*-containing bioaerosols in confirmed TB patients and a majority of randomly selected community members, and we have demonstrated the time-dependent reduction (but not elimination) of *Mtb* bioaerosol positivity in TB clinic attendees irrespective of TB chemotherapy. These observations reinforce the axiom that, while *Mtb* infection is necessary for TB, it is not sufficient: instead, a poorly understood combination of host, environmental, and mycobacterial factors determine disease risk.

In turn, these findings have important implications on the development and testing of new vaccines and prognostic models, as well as the design of future transmission studies and intervention strategies. Although the significance for TB control and the attributable transmission risk from this population remains to be established, these data provide a plausible explanation for the difficulty in linking transmission chains in high-burden regions. Moreover, since the bacilli produced by randomly selected community members appear quantitatively and qualitatively indistinguishable from those released by confirmed TB patients (at least using existing tools to interrogate phenotypic and/or genetic adaptations), it seems plausible that there is some contribution to ongoing *Mtb* transmission. By extrapolating to the broader community, the implication appears unavoidable that there are far greater numbers of disease- and symptom-free individuals with prolonged shedding of *Mtb* in communal settings than those with TB disease.

## Methods

### Study population and participant recruitment

This study was conducted in Masiphumelele, a peri-urban township located south of Cape Town, South Africa. This residential area is divided into demarcated parcels of land, called “erfs” under South African legislation, each with its own unique numeric identifier. Erfs were randomly selected and all consenting participants over 14 years of age were eligible for recruitment into this study. Recruitment was conducted between February–December 2022, with ethical approval from the Human Research Ethics Committee of the University of Cape Town (HREC no. 529/2019).

### The Respiratory Aerosol Sampling Chamber

The RASC is a purpose-built personal clean room equipped with a high efSiciency bioaerosol collection system, which captures all exhaled particulate matter (bioaerosols) at 100–300 L/min in 15 mL of collection medium (sterilized phosphate-buffered saline supplemented with Polymyxin B, Amphotericin B, Nalidixic acid, Trimethoprim and Azilocillin (PANTA) [Becton Dickinson]). Bioaerosol samples were concentrated and stained with DMN-trehalose (Olilux Biosciences Inc.) overnight, as previously reported, before transfer to nanowell devices for imaging (Dinkele et al., 2021).

### Bioaerosol generation and sample processing

This study was conducted in two phases (Fig. 1). In the first phase, participants 1–39 underwent bioaerosol collection during three respiratory maneuvers: FVC, TiBr, and induced cough, as previously described (Dinkele et al., 2022). The sequence of these maneuvers remained consistent throughout the study. During FVC and cough sampling, participants performed the designated maneuver 15 times within a five-minute period, as directed by the study nurse. For tidal breath sampling, participants were instructed to breathe normally into the bioaerosol collection system for five minutes. Each bioaerosol sample underwent independent collection, processing, and enumeration. Enumeration of *Mtb* bacilli was conducted by depositing concentrated and DMN-trehalose-stained bioaerosol samples onto nanowell devices. Bioaerosol positivity was determined by aggregating counts from all three maneuvers. Given the high proportion of bioaerosol positivity within this cohort, all participants were scheduled for a follow-up visit to obtain blood and sputum samples for subsequent QFT and GXP analysis.

The second phase involved the recruitment of participants 40–89 for a longitudinal examination of *Mtb* bioaerosols. This phase comprised 50 individuals sampled on three occasions: baseline, two weeks, and two months. At each visit, two equivalent bioaerosol samples were collected, with each involving ten minutes of tidal breathing interspersed with deep breaths every 30 seconds. Subsequently, these samples were concentrated and arrayed on nanowell devices. Blood and sputum specimens were obtained at baseline for QFT and GXP analysis, respectively.

In all cases, microscopists were blinded to the origin of the sample (which included an empty booth negative control) and enumerated putative *Mtb* bacilli per sample.

### QuantiFERON-TB Gold assay

In this study, BARC SA were contracted to perform and interpret QFT assays according to the manufacturer’s instructions (Qiagen).

### Sputum collection and the GeneXpert assay

For all participants, sputum sampling was attempted. In those capable of producing sputum, a GXP assay was performed and interpreted according to the manufacturer’s instructions (Cepheid). Individuals unable to produce sputum were considered sputum-GXP negative.

### Statistical methods

For the descriptive statistics, a Fisher’s Exact Test was used for the categorical variables given the relatively small sample size. To assess age, the normality and variance of the data were assessed, and a Student’s T Test was used for the comparison. Generalized linear regression was used to assess the bioaerosol results, either as a dichotomous outcome (logistic regression) or a count outcome (negative binomial regression). These outcomes were regressed against age, biological sex, previous TB, QFT results, and HIV status. For the respiratory maneuver comparison, we used the equivalent mixed effects regression models to account for the fact that each participant produced three samples.

## Data Availability

All data produced in the present study are available upon reasonable request to the authors.

## Author contribution

Conceptualization and design: R.D., S.G., B.P., A.M., Z.H., A.V., R.S., A.K., D.F.W. & R.W.; acquisition of data: A.M., Z.H., A.V., R.S; analysis and interpretation: R.D., D.F.W. & R.W.; first manuscript draft: R.D., D.F.W. & R.W.; funding acquisition: D.F.W. & R.W.. All authors critically reviewed and revised the manuscript for intellectual content and approved it prior to submission.

## Acknowledgements

We acknowledge support of the South African Medical Research Council (MRC-RFA-UFSP-01-2013/CCAMP, R.W.) and the National Institute of Allergy and Infectious Diseases of the US NIH (R01AI147347, R.W.). We are grateful for the funding received through the Myco3V TB Research Unit (U19AI162584, R.W & D.F.W.) as well as from the Research Council of Norway (R&D Project 309592, D.F.W.).

The authors have no conflicting interests to declare.

## References

Barry, C. E., Boshoff, H. I., Dartois, V., Dick, T., Ehrt, S., Flynn, J., Schnappinger, D., Wilkinson, R. J. & Young, D. 2009. The spectrum of latent tuberculosis: rethinking the biology and intervention strategies. Nat Rev Micro, 7, 845–855.

Cohen, A., Mathiasen, V. D., Schön, T. & Wejse, C. 2019. The global prevalence of latent tuberculosis: a systematic review and meta-analysis. Eur Respir J, 54.

Coussens, A. K., Zaidi, S. M. A., Allwood, B. W., Dewan, P. K., Gray, G., Kohli, M., Kredo, T., Marais, B. J., Marks, G. B., Martinez, L., Ruhwald, M., Scriba, T. J., Seddon, J. A., Tisile, P., Warner, D. F., Wilkinson, R. J., Esmail, H., Houben, R. M. G. J., Alland, D., Behr, M. A., Beko, B. B., Burhan, E., Churchyard, G., Cobelens, F., Denholm, J. T., Dinkele, R., Ellner, J. J., Fatima, R., Haigh, K. A., Hatherill, M., Horton, K. C., Kendall, E. A., Khan, P. Y., Macpherson, P., Malherbe, S. T., Mave, V., Mendelsohn, S. C., Musvosvi, M., Nemes, E., Penn-Nicholson, A., Ramamurthy, D., Rangaka, M. X., Sahu, S., Schwalb, A., Shah, D. K., Sheerin, D., Simon, D., Steyn, A. J. C., Thu Anh, N., Walzl, G., Weller, C. L., Williams, C. M. L., Wong, E. B., Wood, R., Xie, Y. L. & Yi, S. 2024. Classification of early tuberculosis states to guide research for improved care and prevention: an international Delphi consensus exercise. The Lancet Respiratory Medicine.

Crampin, A. C., Glynn, J. R., Traore, H., Yates, M. D., Mwaungulu, L., Mwenebabu, M., Chaguluka, S. D., Floyd, S., Drobniewski, F. & Fine, P. E. 2006. Tuberculosis transmission attributable to close contacts and HIV status, Malawi. Emerg Infect Dis, 12, 729–35.

Dinkele, R., Gessner, S., Mckerry, A., Leonard, B., Leukes, J., Seldon, R., Warner, D. F. & Wood, R. 2022. Aerosolization of Mycobacterium tuberculosis by Tidal Breathing. American Journal of Respiratory and Critical Care Medicine, 206, 206–216.

Dinkele, R., Gessner, S., Mckerry, A., Leonard, B., Seldon, R., Koch, A. S., Morrow, C., Gqada, M., Kamariza, M., Bertozzi, C. R., Smith, B., Mcloud, C., Kamholz, A., Bryden, W., Call, C., Kaplan, G., Mizrahi, V., Wood, R. & Warner, D. F. 2021. Capture and visualization of live Mycobacterium tuberculosis bacilli from tuberculosis patient bioaerosols. PLOS Pathogens, 17, e1009262.

Dinkele, R., Khan, P. Y. & Warner, D. F. 2024. Mycobacterium tuberculosis transmission: the importance of precision. The Lancet Infectious Diseases.

Dowdy, D. W. & Behr, M. A. 2022. Are we underestimating the annual risk of infection with Mycobacterium tuberculosis in high-burden settings? The Lancet Infectious Diseases, 22, e271–e278.

Drain, P. K., Bajema, K. L., Dowdy, D., Dheda, K., Naidoo, K., Schumacher, S. G., Ma, S., Meermeier, E., Lewinsohn, D. M. & Sherman, D. R. 2018. Incipient and Subclinical Tuberculosis: a Clinical Review of Early Stages and Progression of Infection. Clin Microbiol Rev, 31.

Esmail, H., Dodd, P. J. & Houben, R. M. G. J. 2018. Tuberculosis transmission during the subclinical period: could unrelated cough play a part? The Lancet Respiratory Medicine, 6, 244–246.

Fortune, S. M. 2024. The Titanic question in TB control: Should we worry about the bummock? Proceedings of the National Academy of Sciences, 121, e2403321121.

Glynn, J. R., Guerra-Assunção, J. A., Houben, R. M., Sichali, L., Mzembe, T., Mwaungulu, L. K., Mwaungulu, J. N., Mcnerney, R., Khan, P. & Parkhill, J. 2015. Whole genome sequencing shows a low proportion of tuberculosis disease is attributable to known close contacts in rural Malawi. PloS one, 10, e0132840.

Hermans, S., Horsburgh Jr, C. R. & Wood, R. 2015. A Century of Tuberculosis Epidemiology in the Northern and Southern Hemisphere: The Differential Impact of Control Interventions. PLoS ONE, 10, e0135179.

Horton, K. C., Richards, A. S., Emery, J. C., Esmail, H. & Houben, R. M. 2023. Reevaluating progression and pathways following Mycobacterium tuberculosis infection within the spectrum of tuberculosis. Proceedings of the National Academy of Sciences, 120, e2221186120.

Johnson, G. R. & Morawska, L. 2009. The mechanism of breath aerosol formation. Journal of Aerosol Medicine and Pulmonary Drug Delivery, 22, 229–237.

Jones-LÓpez, E. C., Namugga, O., Mumbowa, F., Ssebidandi, M., Mbabazi, O., Moine, S., Mboowa, G., Fox, M. P., Reilly, N., Ayakaka, I., Kim, S., Okwera, A., Joloba, M. & Fennelly, K. P. 2013. Cough Aerosols of Mycobacterium tuberculosis Predict New Infection. A Household Contact Study. American Journal of Respiratory and Critical Care Medicine, 187, 1007–1015.

Kamariza, M., Shieh, P., Ealand, C. S., Peters, J. S., Chu, B., Rodriguez-Rivera, F. P., Babu Sait, M. R., Treuren, W. V., Martinson, N., Kalscheuer, R., Kana, B. D. & Bertozzi, C. R. 2018. Rapid detection of Mycobacterium tuberculosis in sputum with a solvatochromic trehalose probe. Science Translational Medicine, 10.

Kendall, E. A., Shrestha, S. & Dowdy, D. W. 2021. The Epidemiological Importance of Subclinical Tuberculosis. A Critical Reappraisal. Am J Respir Crit Care Med, 203, 168–174.

Lin, P. L. & Flynn, J. L. 2018. The End of the Binary Era: Revisiting the Spectrum of Tuberculosis. J Immunol, 201, 2541–2548.

Malherbe, S. T., Shenai, S., Ronacher, K., Loxton, A. G., Dolganov, G., Kriel, M., Van, T., Chen, R. Y., Warwick, J., Via, L. E., Song, T., Lee, M., Schoolnik, G., Tromp, G., Alland, D., Barry, C. E., Winter, J., Walzl, G., Lucas, L., Van Der Spuy, G., Stanley, K., Thiart, L., Smith, B., Du Plessis, N., Beltran, C. G. G., Maasdorp, E., Ellmann, A., Choi, H., Joh, J., Dodd, L. E., Allwood, B., Koegelenberg, C., Vorster, M., Griffith-Richards, S. & The Catalysis, T. B. C. 2016. Persisting positron emission tomography lesion activity and Mycobacterium tuberculosis mRNA after tuberculosis cure. Nature Medicine, 22, 1094–1100.

Middelkoop, K., Mathema, B., Myer, L., Shashkina, E., Whitelaw, A., Kaplan, G., Kreiswirth, B., Wood, R. & Bekker, L.-G. 2014. Transmission of Tuberculosis in a South African Community With a High Prevalence of HIV Infection. The Journal of Infectious Diseases, 211, 53–61.

Middelkoop, K., Mathema, B., Myer, L., Shashkina, E., Whitelaw, A., Kaplan, G., Kreiswirth, B., Wood, R. & Bekker, L. G. 2015. Transmission of tuberculosis in a South African community with a high prevalence of HIV infection. J Infect Dis, 211, 53–61.

Mishra, R., Hannebelle, M., Patil, V. P., Dubois, A., Garcia-Mouton, C., Kirsch, G. M., Jan, M., Sharma, K., Guex, N. & Sordet-Dessimoz, J. 2023. Mechanopathology of bioSilm-like Mycobacterium tuberculosis cords. Cell, 186, 5135–5150. e28.

Moyo, S., Ismail, F., Van Der Walt, M., Ismail, N., Mkhondo, N., Dlamini, S., Mthiyane, T., Chikovore, J., Oladimeji, O., Mametja, D., Maribe, P., Seocharan, I., Ximiya, P., Law, I., Tadolini, M., Zuma, K., Manda, S., Sismanidis, C., Pillay, Y. & Mvusi, L. 2022. Prevalence of bacteriologically confirmed pulmonary tuberculosis in South Africa, 2017-19: a multistage, cluster-based, cross-sectional survey. The Lancet Infectious Diseases.

Nguyen, H. V., Tiemersma, E., Nguyen, N. V., Nguyen, H. B. & Cobelens, F. 2023. Disease Transmission by Patients With Subclinical Tuberculosis. Clinical Infectious Diseases.

Pai, M., Dewan, P. K. & Swaminathan, S. 2023. Transforming tuberculosis diagnosis. Nature Microbiology, 8, 756–759.

Patterson, B., Dinkele, R., Gessner, S., Koch, A., Hoosen, Z., January, V., Leonard, B., Mckerry, A., Seldon, R., Vazi, A., Hermans, S., Cobelens, F., Warner, D. F. & Wood, R. 2024. Aerosolization of viable Mycobacterium tuberculosis bacilli by tuberculosis clinic attendees independent of sputum-Xpert Ultra status. Proceedings of the National Academy of Sciences, 121, e2314813121.

Patterson, B., Morrow, C., Singh, V., Moosa, A., Gqada, M., Woodward, J., Mizrahi, V., Bryden, W., Call, C., Patel, S., Warner, D. & Wood, R. 2018. Detection of Mycobacterium tuberculosis bacilli in bio-aerosols from untreated TB patients Gates Open Research, 1.

Richards, A. S., Sossen, B., Emery, J. C., Horton, K. C., Heinsohn, T., Frascella, B., Balzarini, F., Oradini-Alacreu, A., Häcker, B., Odone, A., Mccreesh, N., Grant, A. D., Kranzer, K., Cobelens, F., Esmail, H. & Houben, R. M. G. J. 2023. Quantifying progression and regression across the spectrum of pulmonary tuberculosis: a data synthesis study. The Lancet Global Health, 11, e684–e692.

Shah, N. S., Auld, S. C., Brust, J. C., Mathema, B., Ismail, N., Moodley, P., Mlisana, K., Allana, S., Campbell, A., Mthiyane, T., Morris, N., Mpangase, P., Van Der Meulen, H., Omar, S. V., Brown, T. S., Narechania, A., Shaskina, E., Kapwata, T., Kreiswirth, B. & Gandhi, N. R. 2017. Transmission of Extensively Drug-Resistant Tuberculosis in South Africa. N Engl J Med, 376, 243–253.

Shaikh, N., Swali, P. & Houben, R. M. G. J. 2023. Asymptomatic but infectious – The silent driver of pathogen transmission. A pragmatic review. Epidemics, 44, 100704.

Turner, R. D. & Bothamley, G. H. 2015. Cough and the transmission of tuberculosis. J Infect Dis, 211, 1367–72.

Verver, S., Warren, R. M., Munch, Z., Richardson, M., Van Der Spuy, G. D., Borgdorff, M. W., Behr, M. A., Beyers, N. & Van Helden, P. D. 2004. Proportion of tuberculosis transmission that takes place in households in a high-incidence area. The Lancet, 363, 212–214.

WHO 2023. Global tuberculosis report 2023, World Health Organization.

Williams, C. M., Abdulwhhab, M., Birring, S. S., De Kock, E., Garton, N. J., Townsend, E., Pareek, M., Al-Taie, A., Pan, J., Ganatra, R., Stoltz, A. C., Haldar, P. & Barer, M. R. 2020. Exhaled Mycobacterium tuberculosis output and detection of subclinical disease by face-mask sampling: prospective observational studies. The Lancet Infectious Diseases, 20, 607–617.

Williams, C. M., Muhammad, A. K., Sambou, B., Bojang, A., Jobe, A., Daffeh, G. K., Owolabi, O., Pan, D., Pareek, M., Barer, M. R., Sutherland, J. S. & Haldar, P. 2023. Exhaled Mycobacterium tuberculosis Predicts Incident Infection in Household Contacts. Clin Infect Dis, 76, e957–e964.

Wood, R., Morrow, C., Barry, C. E., 3RD, Bryden, W. A., Call, C. J., Hickey, A. J., Rodes, C. E., Scriba, T. J., Blackburn, J., Issarow, C., Mulder, N., Woodward, J., Moosa, A., Singh, V., Mizrahi, V. & Warner, D. F. 2016. Real-Time Investigation of Tuberculosis Transmission: Developing the Respiratory Aerosol Sampling Chamber (RASC). PLoS One, 11, e0146658.

